# Cascades of effectiveness of new-generation insecticide-treated nets against malaria, from entomological trials to real-life conditions

**DOI:** 10.1101/2025.02.07.25321565

**Authors:** Clara Champagne, Jeanne Lemant, Alphonce Assenga, Ummi A. Kibondo, Ruth G. Lekundayo, Emmanuel Mbuba, Jason Moore, Joseph B. Muganga, Watson S. Ntabaliba, Olukayode G. Odufuwa, Johnson Kyeba Swai, Maria Alexa, Roland Goers, Monica Golumbeanu, Nakul Chitnis, Amanda Ross, Sarah Moore, Emilie Pothin

## Abstract

As insecticide resistance spreads in Africa, new-generation insecticide-treated nets (ITNs) are increasingly being deployed to protect vulnerable populations against malaria. While these nets provide greater entomological efficacy against resistant mosquitoes, their effectiveness against malaria transmission also depends on other factors, such as durability, access, usage, and activity patterns of hosts and vectors. Here, we quantify the impact of two new-generation ITNs, namely Interceptor®G2 (chlorfenapyr-pyrethroid) and Olyset®Plus (piperonyl butoxide-pyrethroid), in a cascade from entomological efficacy to population-level effectiveness. We use a mathematical model that we parameterize with entomological data and validate against results from randomized controlled trials. We found that, beyond entomological factors, operational factors including functional survival, ITN use and in-bed exposure critically impact ITN effectiveness overall and per ITN types. Our results obtained for Tanzania can be extended to other contexts in a dashboard (https://aimswisstph.shinyapps.io/ITNcascadesdashboard) allowing users to explore product selection based on setting-specific factors that influence ITN effectiveness.

## Introduction

Insecticide-treated nets (ITNs) are the cornerstone of malaria control and are recommended by the World Health Organization (WHO) in all malaria-endemic geographies ^1^. Since 2004, over three billion ITNs have been delivered ^2^ and between 2000 and 2022 ITNs have averted an estimated 1.5 billion malaria cases globally ^3,4^. ITNs protect individuals against malaria through three mechanisms. Firstly, they act as a physical barrier, preventing mosquitoes from biting hosts sleeping under the net. Secondly, the insecticide interferes with mosquito host-seeking and prevents bites even when the ITN has become damaged. Thirdly, mosquitoes that get in contact with the net may be killed by the insecticide or suffer non-lethal adverse effects, such as reduced fertility. Therefore, ITNs provide individual protection for the net user and community protection through shorter mosquito survival, reduced mosquito populations and reduced infectious reservoir ^5,6^.

However, malaria vector mosquitoes have become increasingly resistant to public health insecticides, especially on the African continent ^7^. As a result, the insecticidal efficacy of standard ITNs, which contain insecticides from the pyrethroid class such as alpha-cypermethrin, deltamethrin or permethrin, has substantially decreased ^8^. To counteract this effect, new-generation ITNs containing additional active ingredients combined with pyrethroids have been developed. Several have been recommended by the WHO based on randomized controlled trials (RCTs) that demonstrated public health impact ^1^ followed by prequalification ^9^, after their quality, safety and entomological efficacy were demonstrated.

The first class to be pre-qualified contain pyrethroids combined with piperonyl butoxide (PBO), a synergist that restores pyrethroid sensitivity in resistant mosquitoes by inhibiting detoxifying enzymes ^10^. In 2023, the WHO also approved ITNs in which pyrethroids are combined with chlorfenapyr. Chlorfenapyr is a pro-insecticide that becomes active only after being metabolized inside the target insect ^11^. Once ingested or absorbed, it is converted into its active form (tralopyril) by detoxifying enzymes. The active compound disrupts metabolic activity and is effective against mosquitoes that have developed resistance to other insecticides, such as pyrethroids, as it targets a different biological pathway. The improved effectiveness of PBO and chlorfenapyr nets relative to standard pyrethroid ITNs was shown in RCTs and pilot studies ^12–16^ and in 2023, PBO nets represented more than half of the ITNs distributed in sub-Saharan Africa, and dual active ingredient ITNs, which include chlorfenapyr nets, about 20% ^4^.

Unfortunately, insecticide resistance is not the only factor reducing the effectiveness of ITNs. Additional considerations including access to a net, usage of the net by the population and durability of the nets are key to guarantee ITN effectiveness for malaria control ^17^. In 2023, overall access to ITNs was estimated at around 60% in Africa, with only five countries on the continent achieving the target of 80% of the population sleeping under an ITN ^4,18^. Moreover, with national ITN campaigns usually planned to happen every three years, it is crucial that nets are retained and maintain their efficacy over this time frame. In practice, much shorter ITN survivorship has been observed in many settings with an estimated overall median retention time of 1.64 years among all countries in Africa ^18^. Finally, nets only protect hosts when they are under the net and, depending on both mosquito and humans’ activity patterns, a non-negligible proportion of bites may happen when hosts are unprotected while they are not in bed ^19–23^. For nets to be effective at both individual and population level, they have to be present and in regular use, in good physical condition and remain insecticidal, thereby providing protection through preventing bites and killing disease vectors for the population at risk of malaria. Therefore, the effectiveness of ITNs requires consideration of all these factors.

Various indicators exist to monitor insecticide resistance, ITN use, ITN durability and in-bed exposure (see Textbox 1) but these indicators are measured independently of each other and therefore do not provide information on the relative contribution of each factor to the overall public health impact of ITNs. Quantifying this relative contribution is however of public health relevance because it allows stakeholders to identify context specific factors that reduce the impact of ITNs, and therefore to prioritize strategies to address them.

The contribution of these different factors on the pathway from efficacy to effectiveness can be explored using the concept of effectiveness cascade ^24^. Such cascades have often been used to represent the impact of health systems factors on malaria case management, both conceptually ^25–27^ and quantitatively ^28,29^. For vector control and ITNs in particular, while conceptual cascades have been presented in the literature ^24,30^, they have rarely been formally quantified ^31–33^ due to the difficulty of disentangling factors that are intrinsically intertwined in real-life settings. Mathematical models of malaria transmission offer the possibility to disentangle each factor and quantify their relative contribution to the overall ITN effectiveness ^34^.

With mathematical models of malaria transmission, epidemiological conditions can be reproduced *in silico* and the impact of specific control interventions on risk and burden can be quantified in areas or configurations in which they have not formally been tested through trials ^35–40^. Therefore, these models enable us to extract further information from the entomological and epidemiological trials, once they have been calibrated and validated against the data from these trials. In particular, EHT data has been used to estimate the entomological efficacy of various vector control tools and hence parameterize mathematical models of malaria ^41–44^. Population-level RCTs can be used as out-of-sample validation sets to evaluate the capacity of the model to predict epidemiological outcomes accurately ^39,40,44^. Regarding ITNs specifically, parameterized and validated models have been developed for pyrethroid and PBO ITNs^41,44^ as well as chlorfenapyr ITNs^39^, but they have not been used to quantify cascades of LLIN effectiveness.

In this work, we quantify the epidemiological impact of two new-generation ITNs, namely Olyset® Plus (PBO-pyrethroid) and Interceptor® G2 (chlorfenapyr-pyrethroid) in Tanzania with an established mathematical model of malaria transmission, the OpenMalaria modelling suite ^45^. The model is calibrated to five EHT entomological datasets and validated against two RCTs from the same country for prevalence outcomes. The model is then used to quantify the impact of both net types along the different steps of the effectiveness cascade and identify the main factors responsible for the loss of effectiveness.

## Texbox 1: Definitions

### Entomological Efficacy

“as measured by a reduction in vectorial capacity in experimental huts indicates that the intervention (e.g., ITNs) is effective in reducing the potential of mosquitoes to act as disease vectors, by decreasing their survival rates, deterring biting, or limiting their capacity to complete feeding cycles. This measure estimates the change in the likelihood of disease transmission in a defined area if an intervention was deployed at scale”^46^.

### ITN use

Proportion of the population sleeping under a net the previous night. It is the result of both “access” to a net (proportion of the population with access to an ITN for every two persons that slept in the household the previous night) and “use given ownership” (proportion of the population with ITNs that slept under a net the previous night).

### Insecticidal durability

**“**Retention of insecticidal efficacy of the ITN, i.e. content of insecticide meets required standards when measured in laboratory tests and/or sufficient insecticidal availability is demonstrated on the surface of the net by mosquito bioassays. Insecticidal persistence can therefore be broken down into two aspects: persistence of insecticidal content and persisting bio-availability of insecticide. *Synonyms: insecticidal persistence, chemical durability, insecticidal life-span”* ^46^.

### Functional survival

“An estimate of the physical life span of an ITN product in the field, either a general estimate or tailored for specific conditions. It is measured as an ITN that is present in use and in serviceable condition” ^46^.

### Serviceable condition

“an ITN with a proportionate hole index in the ‘good’ (pHI 0-64) or ‘acceptable’ (pHI65-642) range. Any net with a pHI higher than this is termed unserviceable and ‘torn’, the “protective efficacy for the user (is) in serious doubt and the net should be replaced urgently” ^46^.

### In-bed exposure

“The proportion of vector bites occurring indoors during sleeping hours, for an unprotected individual, which represents the maximum possible personal protection any intervention targeting sleeping spaces could provide”^47^.

## Results

### Estimation of new-generation ITN entomological efficacy from experimental hut trial data

Entomological efficacy was quantified in four indicators: reduction in host availability (reflecting the deterrent properties of the nets), pre-prandial killing effect (referring to mosquito mortality before biting), post-prandial killing effect (referring to mosquito mortality after biting) and overall reduction in vectorial capacity (representing to the potential for a given mosquito population to transmit malaria by combining the previous three indicators). The entomological efficacy of Olyset® Plus and Interceptor® G2 was compared to pyrethroid-only ITNs, estimated using five EHTs conducted in Tanzania referred to here as BIT103^48^, BIT055, BIT080^49^, Odufuwa^50^, and Kibondo ^51^ (cf. Figure 1, with all numerical outputs in Supplementary Table 1). Over all trials, the reduction in host availability was estimated to be higher for Olyset® Plus compared to Interceptor® G2. On the contrary, the increase in host-seeking mortality and the pre-prandial killing effect were higher for Interceptor® G2, except for the Odufuwa^50^ trial which compared Olyset Plus to PermaNet 2.0. Post-prandial killing effects were more uncertain and displayed stronger variations across trials: this can be explained because very few fed mosquitoes were collected in all trials (see Appendix 4), thus the proportion of killed among fed mosquitoes was estimated from very few observations.

**Figure 1:**
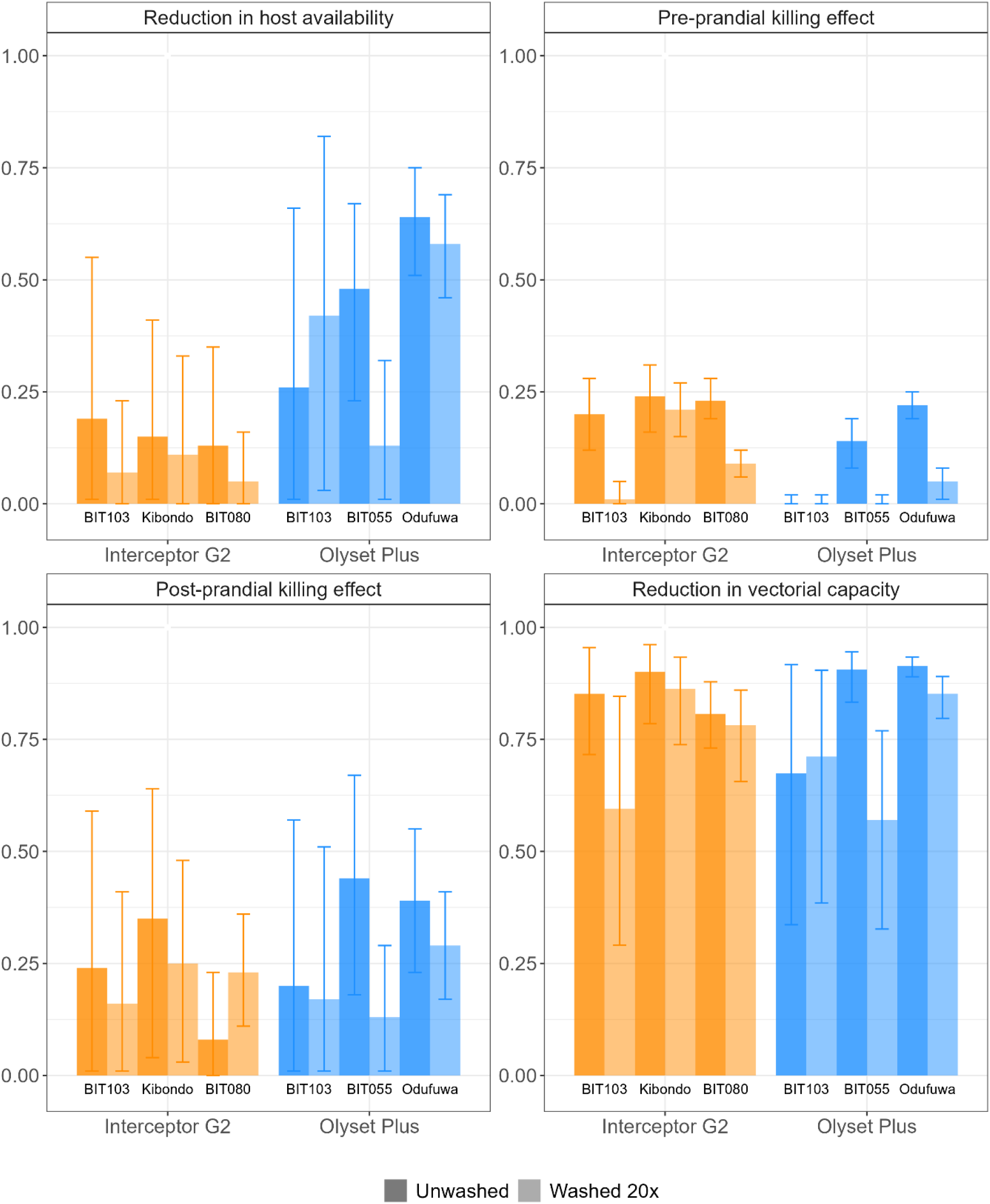
Estimates of entomological efficacy for unwashed and washed nets (posterior mean and 95% credible intervals), compared to pyrethroid-only nets as control. The various experimental huts considered are BIT103^48^, BIT055, Odufuwa et al.^50^, BIT080^49^ and Kibondo et al. ^51^. The active ingredient in pyrethroid-only controls is alpha-cypermethrin in BIT103^48^, BIT080^49^ and Kibondo et al. ^51^, permethrin in BIT055 and deltamethrin in Odufuwa et al.^50^.

Combining all three indicators in the vectorial capacity, the overall entomological efficacy of Interceptor® G2 was high (above 80%) in the three trials. For Olyset® Plus, the reduction of vectorial capacity was high in the Odufuwa^50^ and BIT055 trials (above 90%) but lower in BIT103 (67%). These three trials used different pyrethroid-only nets as control (deltamethrin in Odufuwa et al.^50^, permethrin in BIT055, and alpha-cypermethrin in BIT103), and this could explain some of these differences. In the data from the BIT103 trial, which includes both new-generation net types within the same trial, Interceptor® G2 was estimated to have a higher efficacy than Olyset® Plus (reduction in vectorial capacity of 85% (72-95) Interceptor® G2 compared to 67% (34-92) for Olyset® Plus).

All three EHTs include arms in which ITNs have been washed 20 times, in order to represent what the efficacy due to insecticide content would be after three years of operational use ^52^. All indicators were lower for washed nets compared to unwashed nets, except in two cases. In BIT103 for Olyset® Plus, repellency and overall vectorial capacity reduction were higher for washed nets than unwashed nets, although this result needs to be interpreted with caution given the very large uncertainty around the point estimate. In BIT080, washed Interceptor® G2 nets were estimated to have a higher post-prandial killing efficacy but a lower pre-prandial killing efficacy than unwashed ones, and overall, vectorial capacity was slightly lower for the washed nets. Overall, the decrease in entomological efficacy due to washing was small in most trials, except in BIT103 for Interceptor® G2 and BIT055 for Olyset® Plus.

### Model validation: reproduction *in silico* of randomized control trial data

The estimates of entomological efficacy, alongside setting-specific information related to epidemiological conditions and ITN effectiveness, were inputted in a mathematical model of malaria transmission, the OpenMalaria simulation platform ^45^. To assess the capacity of the model to quantify the epidemiological impact of ITN deployments, two randomized controlled trials conducted in Tanzania, Mosha et al. ^12,13^ and Protopopoff et al. ^53,54^ were reproduced *in silico* (cf. Figure 2). The model was adjusted to reproduce the control arm (with pyrethroid-only nets) and the pre-intervention prevalence in the intervention arms (with Olyset® Plus and Interceptor® G2 nets), while the post-intervention prevalence estimates in the intervention arms were used for validation only. For both trials, the uncertainty intervals for the model predictions and observations overlapped at all time points but one. In the trial by Mosha et al.^12,13^, the observation at 24 months in all arms was slightly missed. In the trial by Protopopoff et al. ^53,54^, the observation at 28 months in the intervention arm was largely missed; however, the observed prevalence at that date was abnormally high in both the control and intervention arm: the estimates being higher than all other measurements including the pre-intervention estimate. Overall, the correlation between predictions and observations was 0.85 for prevalence and 0.80 for effect sizes (relative difference between prevalence in the intervention arm and the control) and the R^2^ are 0.72 and 0.63 respectively (scatterplots are displayed in Appendix 4).

**Figure 2.**
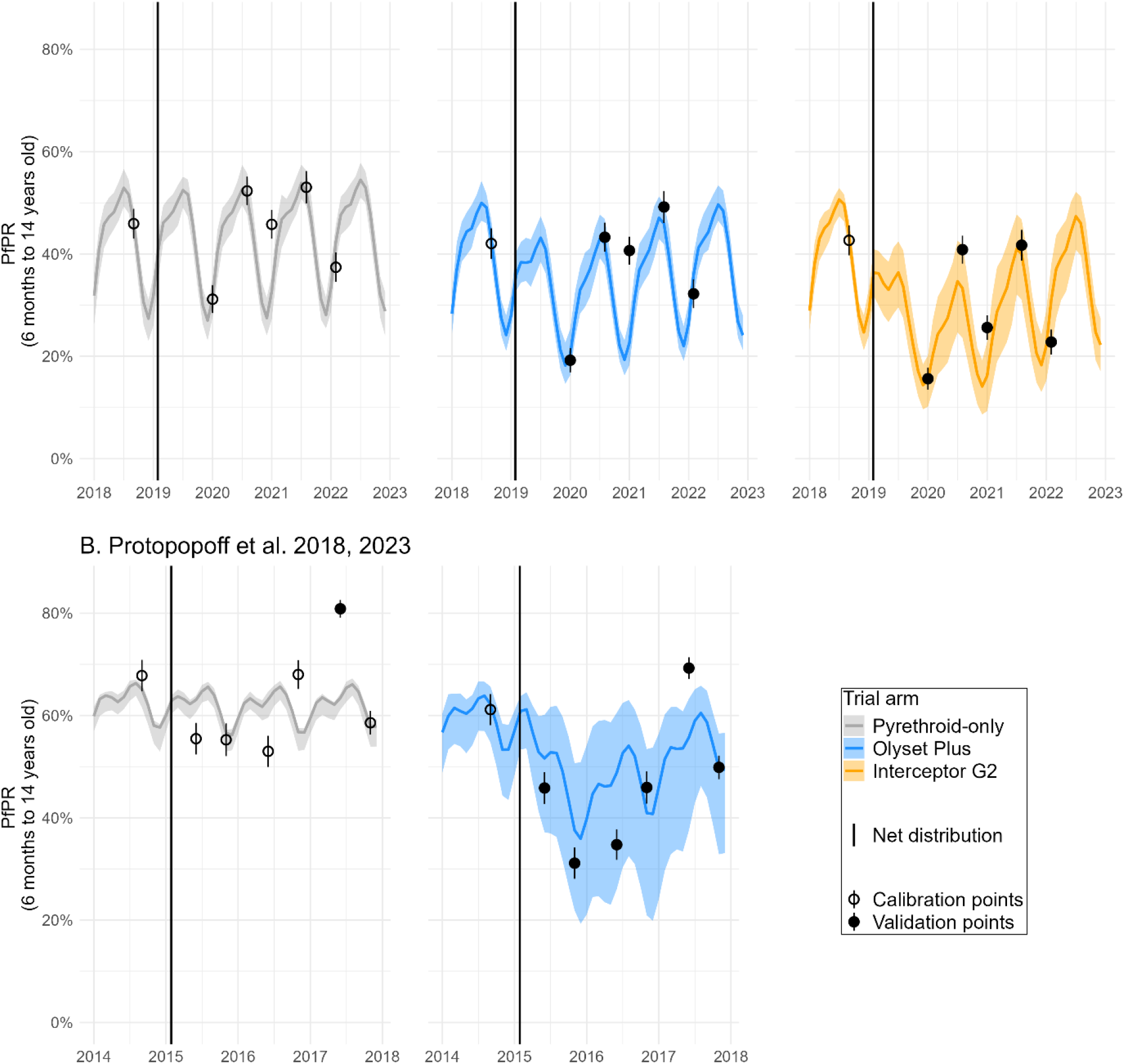
Validation of model Plasmodium falciparum prevalence (PfPR) estimates (curve and shaded areas) against measurements from the RCTs (dots). Plain dots indicate observations that were not used to calibrate the model and are solely presented for validation. White dots were used in the calibration process. Plain line represent the mean model prediction, while shaded areas represent model uncertainty associated with EHT fitting, transmission intensity calibration and model stochasticity (see Methods section for methodological details). A.Trial by Mosha et al.^12,13^ B. Trial by Protopopoff et al.^53,54^.

#### Cascades of ITN effectiveness

Finally, we used the validated model to measure the decline in effectiveness between entomological efficacy and population-level conditions, and quantified the relative contribution of five factors to this decline: entomological efficacy, functional survival, usage at distribution, insecticidal survival and in-bed exposure (Figure 3). Effectiveness was measured in terms of vectorial capacity reduction, and population-level conditions are taken to represent the trial by Mosha et al.^12,13^, but assumptions can be modified to reflect other settings in an interactive dashboard: https://aimswisstph.shinyapps.io/ITNcascadesdashboard.

**Figure 3.**
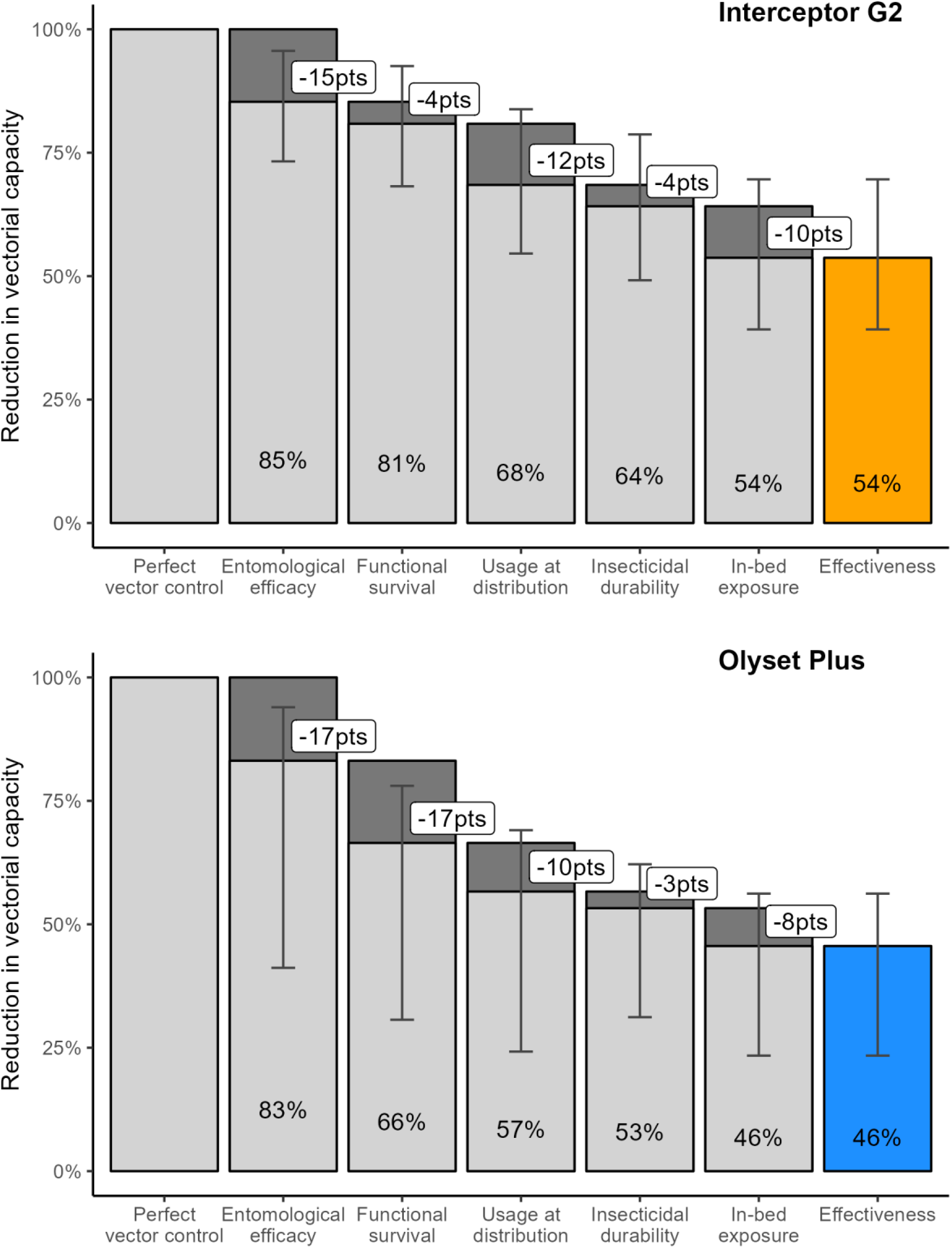
Cascades of ITN effectiveness, reflecting average entomological efficacy estimates from all five EHT and setting-specific conditions of the Mosha et al. ^12,13^ randomized controlled trial. Top panel (orange): Interceptor G2. Bottom panel (blue): Olyset Plus. Bars represent the mean and error bars represent the 95% credible intervals from EHT fitting of entomological efficacy, based on 1000 samples from each EHT and taking the ensemble over all EHT for each net type (see Methods section). Cascades for each EHT individually are displayed in Appendix 1.

While the effectiveness of both net types was above 80% when only entomological efficacy was considered, it dropped below 55% when all other dimensions were included, highlighting the large contribution of operational factors beyond the sole killing and repelling properties of the ITNs.

The overall effectiveness of Interceptor G2 nets was higher than the one of Olyset Plus nets, reducing vectorial capacity on average by 54% over three years, against 46% for Olyset Plus. The largest difference in effectiveness between the two ITNs came from functional survival: this factor was responsible on average for a drop of 4 percentage points for the Interceptor G2 against 16 for the Olyset Plus. On the contrary, the overall differences in entomological efficacy between the two ITNs were small when averaging across EHT (cascades per EHT are displayed in Appendix 1 and in the interactive dashboard).

Beyond entomological efficacy, the main factors responsible for the decline in effectiveness for both ITNs were imperfect usage at distribution (responsible for a drop of 10 to 12 points) and in-bed exposure (responsible for a drop of 8 to 10 points). On the other hand, insecticidal durability had a smaller role in the loss of effectiveness (responsible for a drop of 3 to 4 points).

## Discussion

In this work, we quantify the impact of two new-generation ITNs, namely Interceptor G2 (chlorfenapyr-pyrethroid) and Olyset Plus (PBO-pyrethroid), in a cascade from entomological efficacy to population-level effectiveness. We found that, beyond entomological factors, operational factors such as functional survival, initial ITN use and in-bed exposure play a crucial role in ITN impact, both overall and when comparing ITN types. Our results obtained for Tanzania can be extended to other settings thanks to an interactive dashboard (https://aimswisstph.shinyapps.io/ITNcascadesdashboard) allowing users to explore various combinations of activity rhythms, durability properties, ITN use and mosquito species.

These cascades make it possible to explore the main bottlenecks reducing the effectiveness of ITNs at the population level. During the RCT conducted by Mosha et al. ^12,13^, functional survival was highlighted as a substantial contributor to effectiveness decay for Olyset Plus, while imperfect usage and exposure outside sleeping hours were important for both net types. Such considerations provide valuable information for decision-makers who face resource-constrained choices such as transitioning to new-generation nets, improving deployment strategies, improving ITN access between mass distribution campaigns, or introducing complementary outdoor vector-control interventions. Although all these interventions are valuable, understanding their relative importance can help focus efforts on the most critical factors for maximizing malaria control in a given setting.

Our model is based on estimates of entomological efficacy inferred from five experimental hut trials conducted in Tanzania. We found that Interceptor G2 presents strong pre-prandial killing properties while Olyset Plus has a higher deterrence effect when compared to pyrethroid-only nets as control. Substantial variation was observed across experimental hut trials, which can likely be explained by the type of comparator pyrethroid net used (permethrin, deltamethrin or alpha-cypermethrin). In entomological trials the choice of active comparator is a critical determinant of relative ITN performance. It is often assumed that all pyrethroids behave in the same way, but there is abundant evidence that their mode of action varies from quite high deterrence to very low deterrence ^55^. Additionally, EHT data often present large intrinsic variability in mosquito numbers and mortality^10,56^ that could contribute to these variations. Our statistical model to infer entomological efficacy estimates from EHT data is general and could be readily applied to other EHT datasets beyond the five studies presented here, to better understand the drivers of entomological efficacy and the variations between trials^10,56,57^.

At the population level, our model is able to reproduce the reduction in malaria prevalence observed in two RCTs conducted in Tanzania. Therefore, this validated ITN model can now be used for subnational tailoring of interventions and/or country-wide predictions of the impact of malaria interventions ^37,58^. The Tanzanian RCTs reported a substantial effectiveness of new-generation ITNs compared to pyrethroid-only ones, although with substantial differences in duration across net types and trials (3 years for Interceptor G2 in ^12,13^ and Olyset Plus in ^53,54^ versus 1 year for Olyset Plus in ^12,13^). This evidence using Tanzania data is in line with the results observed in other geographies. An RCT comparing PBO vs standard ITNs in Uganda found a superior effect of the PBO nets up to 18 months post distribution and while also highlighting rapid loss of ITNs ^59^. A RCT comparing chlorfenapyr vs standard ITNs in Benin found a superior effect of the chlorfenapyr nets over the first two years after distribution ^14^. The effectiveness pilots conducted in Burkina Faso, Mozambique, and Rwanda also found a superiority of chlorfenapyr and PBO nets against pyrethroid ones over at least two years after distribution. Less clear results were observed in Nigeria where diverging trends in prevalence and incidence could be attributed to data quality issues, confounding interventions and external factors, and where ITNs usage was low^16^.

Our mathematical model sheds light on the differences in impact between the entomological and the population levels, by identifying the main contributors of ITN effectiveness beyond entomological efficacy. Functional survival was highlighted as the main factor responsible for the loss of effectiveness of the studied PBO net in Mosha et al. ^12,13^. This was not so pronounced for Interceptor G2 in that trial, but ITN survival can vary widely across countries due to differences in the net use environment and user behaviour ^34^. A large decrease in Interceptor G2 use in the third year post-distribution was observed in another trial conducted in Benin that was sufficient to reduce impact on malaria^15^. Functional survival is dependent on both human behaviour and fabric integrity as nets are discarded by their users when they are perceived as too torn even if they are still effective in killing mosquitoes ^60^. We estimated functional survival from survey data on ITN usage among trial participants but longitudinal follow-up of the nets in the same setting revealed lower functional survival estimates ^61^. In RCTs, there are frequent hang up, keep up and community sensitizations that maximize ITN access and use. In real conditions, populations receive new ITNs every three years or less without supplemental ITNs or regular net care messaging, so functional survival is expected to be lower ^18^ and would therefore have an even higher contribution to the loss of effectiveness.

ITN use and access at the time of distribution was also identified as a key contributor to ITN impact, despite the high values observed in the RCTs (around 70% or above). These high values are very likely due to the controlled nature of the RCT, where access to a net is not an issue and sensitization during the trial may help ensure a high usage given ownership. In real settings, however, the proportion of people sleeping under a net could be lower even just after distribution, strongly driven by limited access to ITNs ^18^, and would thus also act as a major bottleneck for ITN effectiveness.

Activity rhythms of humans and mosquitoes were also highlighted as a substantial contributor of ITN effectiveness. This is specific to the Tanzanian dataset on human and mosquito activity patterns used in the model, in which human individuals wake up very early, while mosquitoes are still active. Additional representative data on vector and population behaviours would be important to further quantify this impact^19,22^. Overall, this aspect should not be overlooked, and tools that address human-vector contact outside of sleeping hours may have a role to play in reducing the gaps in the effectiveness cascade.

Even though our model has been validated to reproduce controlled settings in Tanzania, the predicted effectiveness cascade in other contexts can be explored with our interactive dashboard. Ecological or behavioural characteristics, such as mosquito species and activity rhythms can be modified to represent other geographies. Intervention characteristics, such as ITN access or usage and durability (median functional survival), can also be modified to represent operational settings beyond the optimal deployment conditions observed in RCTs. Therefore this online interactive tool can enable malaria programs and stakeholders to explore the cascades using setting-specific assumptions relevant to their context.

This work has nonetheless some limitations. First, we only estimated the entomological impact of new-generation nets compared to pyrethroid-only nets as the standard of care, and not compared to the absence of intervention. For this reason, in the RCT, we did not validate the predictions in the control arm, but rather adjusted the model such that this arm was best reproduced. In our adjustment, the effect of pyrethroid-only nets in the control arm was estimated to be very small. This is in agreement with the observations from the Tanzanian RCT used in this work, but in other settings, it was seen to be higher ^59^. Therefore, while this approach does not enable quantification or validation of the effectiveness of pyrethroid-only nets, it has the advantage of highlighting that pyrethroid ITNs vary in their mode of action and careful selection of pyrethroid only ITNs in experimental hut trials is needed. Second, due to our constrained calibration method, we did not have enough flexibility to perfectly reproduce the control arm with pyrethroid-only ITNs in the trial by Protopopoff et al. ^53,54^. Previous modelling attempts to reproduce this trial with another malaria model faced very similar issues and outcomes ^44^. In particular, in our case, one observation from ^54^, with a reported prevalence above 80% among children under 15 years old, was treated as an outlier. It was displayed as a validation point not used during calibration, considering that the occurrence of such high prevalence in the trial is likely due to external factors not included in the model. Third, our estimates of post-prandial killing efficacy of the different net types display large uncertainties, due to small sample sizes among fed mosquitoes in this setting. These uncertainties are nonetheless quantified through our statistical model and propagated into the mathematical modelling framework and effectiveness cascades. Fourth, we quantified insecticidal durability comparing unwashed and 20-times washed nets, although there is recent evidence that washing may not realistically demonstrate insecticide loss under user conditions for PBO or chlorfenapyr nets^62,63^: the effect of insecticidal durability on overall effectiveness may therefore be underestimated. Finally, due to the controlled nature of the RCT, some elements influencing ITN effectiveness are not included in our cascade, such as seasonal ITN use ^64^. Nevertheless, provided that precise data on these quantities is available, the framework could be extended to include them.

In conclusion, we have quantified the loss of ITN impact along a cascade from entomological efficacy to population-level effectiveness, highlighting the importance of operational factors such as functional survival, ITN use or in-bed exposure. The performance of an ITN is dependent on several components beyond the insecticide that result in substantial differences in effectiveness. As these differences may be setting-specific due to different mosquito profiles, human behaviour, environment and net care behaviour, our framework allows the combination of multiple data sets to better understand essential components of product effectiveness and enables setting-specific selection of ITN products for use in malaria control and elimination.

## Methods

### Estimating ITN entomological efficacy from experimental hut trial data

#### Experimental hut trial data

Data from five experimental hut trials (EHT) are used:

- Kibondo et al. ^51^ compared chlorfenapyr-pyrethroid nets (Interceptor G2) with alpha-cypermethrin nets (Interceptor)
- BIT080^49^ compared chlorfenapyr-pyrethroid nets (Interceptor G2) with alpha-cypermethrin nets (MiraNet)
- BIT103^48^ compared chlorfenapyr-treated nets (Interceptor G2) and PBO-pyrethroid nets (Olyset Plus) with alpha-cypermethrin nets (MAGNet)
- BIT055 compared PBO-pyrethroid nets (Olyset Plus) with permethrin nets (Olyset)
- Odufuwa et al. ^50^ compared PBO-pyrethroid nets (Olyset Plus) with deltamethrin nets (PermaNet 2.0)

In all trials, the pyrethroid-only nets are taken as the control. Each trial reports the number of mosquitoes found unfed-alive, unfed-dead, fed-alive and fed-dead immediately after collection and then held in a temperature-controlled facility to assess delayed mortality at 24 hours (all trials) and 72 hours (all trials except BIT055 and Odufuwa et al.^50^). This information is summarized over all nights for each experiment in Appendix 4. All analyses are restricted to *Anopheles arabiensis* (due to low total numbers collected for other *Anopheles* species). For each trial, the main analysis used the longest holding time to assess mortality (72 hours for Kibondo et al. ^51^, BIT080 and BIT103 and 24 hours for BIT055 and Odufuwa et al.^50^). As a robustness check, the analysis using 24 hours for all trials is presented in Appendix 2.

#### Statistical model

The reduction in host availability (deterrence), the pre-prandial killing effect, and the post-prandial killing effect are estimated using a Bayesian hierarchical model, adapted from Denz et al. ^42^. In this model, each night *t*, and in each trial arm *i*, mosquitoes have the possibility to feed with rate *α*_*i,t*_ or die during host-seeking with rate *μ*_*i,t*_. Those who have fed have then a probability 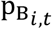 to survive after feeding. At the end of the night, mosquitoes are found in the four categories “unfed-alive” (UA_*i,t*_), “unfed-dead” (UD_*i,t*_), “fed-alive” (FA_*i,t*_ and “fed-dead” (FD_*i,t*_), with respective probabilities 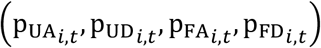, with 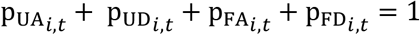. The model can thus be written with the following equations (all notations are indicated inTable 1):

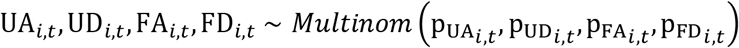

With

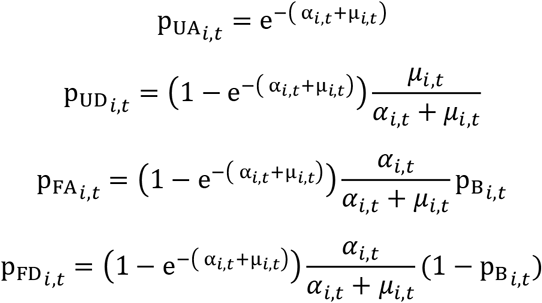

**Table 1.** Parameter definitions and associated prior distributions. Prior distributions are from ^42^.

| QUANTITY | DEFINITION | RANGE/PRIOR | UNIT |
| --- | --- | --- | --- |
| **DATA** |  |  |  |
| $UA_{i,t}$ | Number of mosquitoes unfed and alive at the end of night $t$ in arm $i$ | $\geq 0$ | dimensionless |
| $UD_{i,t}$ | Number of mosquitoes unfed and dead at the end of night $t$ in arm $i$ | $\geq 0$ | dimensionless |
| $FA_{i,t}$ | Number of mosquitoes fed and alive at the end of night $t$ in arm $i$ | $\geq 0$ | dimensionless |
| $FD_{i,t}$ | Number of mosquitoes fed and dead at the end of night $t$ in arm $i$ | $\geq 0$ | dimensionless |
| $F_{i,t}$ | Number of mosquitoes fed at the end of night $t$ in arm $i$ ($FA_{i,t} + FD_{i,t}$) | $\geq 0$ | dimensionless |
| **FITTED PARAMETERS** |  |  |  |
| $\pi_i$ | Reduction in host availability ($\pi_0 = 0$) | Lognormal(0,5) on $1 - \pi_i$ | dimensionless |
| $\kappa_i$ | Increase in the killing rate while the mosquito is host-seeking ($\kappa_0 = 0$) | Lognormal(0,5) | dimensionless |
| $\xi_i$ | Post-prandial killing effect ($\xi_0 = 0$) | Uniform(0,1) | dimensionless |

| HYPER PARAMETERS (FITTED) |  |  |  |
| --- | --- | --- | --- |
| **a, m** | Mean of $\log(\alpha_{0,t})$ and $\log(\mu_{0,t})$ | $N(0,6)$ | $\text{time}^{-1}$ |
| **b** | Mean of $\text{logit}(p_{B,0,t})$ | $\text{Logistic}(0,1)$ | dimensionless |
| TRANSFORMED QUANTITIES |  |  |  |
| **$p_{A,i,t}$** | Probability to remain in the host-seeking stage | $[0,1]$ | dimensionless |
| **$p_{F,i,t}$** | Probability of feeding | $[0,1]$ | dimensionless |
| **$p_{M,i,t}$** | Probability to die while host-seeking | $[0,1]$ | dimensionless |
| **$p_{B,i,t}$** | Probability to survive after feeding | $[0,1]$ | dimensionless |
| **$\alpha_{i,t}$** | Feeding rate | $\geq 0$ | $\text{time}^{-1}$ |
| **$\mu_{i,t}$** | Death rate while host seeking | $\geq 0$ | $\text{time}^{-1}$ |
| **$\phi_i$** | Pre-prandial killing effect | $[0,1]$ | dimensionless |

**Table 2.** Trial-specific assumptions for each of the 3 considered RCTs.

| Parameter | Assumptions and sources | Mosha et al. <sup>12,13</sup> | Protopopoff et al. <sup>53,54</sup> |
| --- | --- | --- | --- |
| **Demography and age structure** | Derived from Ekström et al. <sup>67</sup> | Tanzania |  |
| **Mosquito species** | From trial measurements | *An. funestus* (95%) and *An. gambiae* (5%) <sup>12</sup> | *An. arabiensis* (4%), *An. funestus* (4%) and *An. gambiae* (92%) <sup>53</sup> |
| **Mosquito bionomics** | Species-specific estimates <sup>21</sup>. |  |  |
| **In-bed exposure** | Methodology by Golumbeanu et al. <sup>21</sup>, averaging data from the several studies, as detailed in Appendix 6 | The coefficient representing exposure in bed is estimated to be $\epsilon = 0.69$. | |
| **Seasonality profile** | Based on rainfall data, extracted from CHIRPS via the R package *chirps* <sup>68</sup> | for the Misungwi council (Mwanza region) for the period 2018-2022 | for the Muleba council (Kagera region) for the period 2014-2017 |
| **Case management effective coverage** | Calculated following Galactionova et al. <sup>28</sup> | 2017 DHS survey for Mwanza province (61.6% effective coverage) | 2017 DHS survey for Kagera province (37.8% effective coverage) |
| **ITN usage pre-intervention** | From trial measurements | 59.6% for the pyrethroid arm, 59.3% for the Interceptor G2 arm and 61.7% for the Olyset Plus arm <sup>12</sup> | 30% for the pyrethroid arm and 26% for the Olyset Plus arm <sup>53</sup> |
| **ITN functional survival half-life** | Inferred from the usage measured in the trial, using a Weibull function (see Appendix 5) | Estimated to 2.3 years for the pyrethroid arm, 2.4 years for the Interceptor G2 arm and 1.7 years for the Olyset Plus arm | Estimated to 2.7 for the pyrethroid arm and 2.4 years for the Olyset Plus arm |
| **ITN usage post-intervention** | Inferred from the measured usage in the trial, using a Weibull function (see Appendix 5) | Estimated to 77% for the pyrethroid arm, 69% for the Interceptor | Estimated to 77% for the pyrethroid arm and 81% for the Olyset Plus arm |
| at deployment |  | G2 arm and 75% for the Olyset Plus arm |  |
| Entomological inoculation rate in the absence of interventions (EIR) | Calibrated to reproduce pre-intervention prevalence from RCTs, using maximum likelihood to find the point estimate and profile likelihood methods<sup>69</sup> to infer uncertainty ranges as detailed in Appendix 7. | Estimated to 50 (40-60) for the pyrethroid arm, 40 (34-48) for the Interceptor G2 arm and 38 (32-46) for the Olyset Plus arm. | Estimated to 200 (134-200) for the pyrethroid arm and 98 (46-176) for the Olyset Plus arm |

The effect of a given intervention is measured by comparing the rates *α*_*i,t*_ (feeding rate) and *μ*_*i,t*_ (mortality rate while host-seeking) and the survival probability after biting 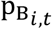 between the intervention arm (chlorfenapyr or PBO net) and the control arm (unwashed pyrethroid-only nets, denoted with *i* = 0).

The parameter *π*_*i*_ corresponds to the reduction in host availability for protected hosts in intervention arm *i* and thus represents the deterrence effect. It is defined as follows:

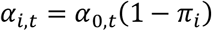

When *π*_*i*_ = 0, the intervention has no deterrence effect, and when *π*_*i*_ = 1, protected hosts escape mosquito bites entirely.

The parameter *κ*_*i*_ corresponds to an increase in the killing rate while the mosquito is host-seeking in intervention arm *i* and it is defined as follows:

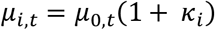

When *k*_*i*_ = 0, the intervention has no effect on host-seeking mortality, and when *κ*_*i*_ > 0 mosquitoes have a higher mortality while host-seeking than in the control. A related indicator is the pre-prandial killing effect *ϕ*_*i*_, namely the reduction in the probability 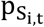 of surviving before biting (where 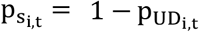. The pre-prandial killing effect can be obtained via the following formula (which can be sampled while fitting the Bayesian model):

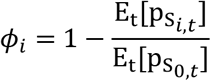

This can be rewritten as

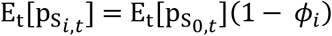

When *ϕ*_*i*_ = 0, the intervention has no effect on the probability that mosquitoes survive, and when *ϕ*_*i*_ = 1, all mosquitoes die and none can feed the protected host. This indicator thus depends on both *π*_*i*_ and *κ*_*i*_ but it is required for subsequent mathematical modelling with OpenMalaria and will be used in the model to represent the potential of the intervention to kill mosquitoes before feeding.

Finally, the parameter *ξ*_*i*_ corresponds to the post-prandial killing effect in intervention arm *i*, whereby the probability of mosquito survival after biting is decreased by the intervention.

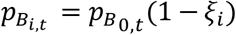

When *ξ*_*i*_ = 0, the intervention has no effect on post-prandial survival, and when *ξ*_*i*_ = 1 no mosquito survives after biting when in contact with the intervention.

The model is fitted with Stan using the Rstan interface ^65^, using 3 MCMC chains with 6000 iterations each, including 3000 warm-ups (10,000 iterations and 7,000 warm-ups for the washed nets in BIT103). Prior distributions and definition ranges for each parameter are indicated in Table 1. The model is fitted independently for each EHT, and for unwashed and 20 times washed nets separately (taking unwashed pyrethroid-only nets as the control in both cases). Convergence diagnostic plots are presented in Appendix 8.

#### Vectorial capacity reduction

The vectorial capacity is defined as the total number of infectious mosquito bites originating from each mosquito biting an infected human. In the model by Chitnis et al. ^66^, which is the non-periodic analog of the model used in OpenMalaria, the vectorial capacity can be calculated either in the absence or in the presence of ITNs in the population to quantify the reduction in vectorial capacity attributable to the ITN intervention ^21^. A perfect vector control tool that would entirely stop malaria transmission corresponds to a 100% reduction in vectorial capacity, while a completely ineffective intervention corresponds to a 0% reduction.

In order to account for the decay in ITN effectiveness over time, the model’s equilibrium vectorial capacity is calculated for different time steps over a 3-year period. At each time step, effective coverage is updated to reflect functional survival and entomological efficacy is updated to reflect insecticidal durability. The reductions in vectorial capacity calculated for each time step are averaged to obtain a single summary value over the 3-year time frame.

In order to propagate the fitting uncertainty of the entomological efficacy estimates, vectorial capacity is calculated for 1000 random samples from the posterior distribution for each EHT. The posterior mean and the 95% credible intervals in the vectorial capacity estimates for each EHT were selected. Vectorial capacity computation is deterministic and the only source of uncertainty propagated is the posterior estimation of the reduction in host availability and the pre- and post-prandial killing effects.

In order to reflect entomological efficacy only (in Figure 1), vectorial capacity is computed under ideal conditions, namely ITN usage is assumed to be 100%, functional survival is assumed to be 3 years, insecticide decay is assumed to be absent and in-bed exposure is assumed to be 100% (cf. perfect comparator in Table 3). Other conditions are explored subsequently in the cascade of effectiveness and the associated methodology is detailed below.

**Table 3.** Realistic estimates and perfect comparator used in the effectiveness cascades to represent the setting in Mosha et al. 2022; 2024).

|  | **Realistic estimates to represent the setting in Mosha et al.** <sup>12,13</sup> | **Perfect comparator** |
| --- | --- | --- |
| **Entomological efficacy** | Deterrence, pre-prandial and post-prandial killing effects as fitted from the EHT data, combined with a standard pyrethroid-only net with 0.05 pre-prandial killing effect. | Assuming a 100% reduction in vectorial capacity (perfect vector control tool) |
| **Functional survival** | Weibull decay function with half-life and shape parameters fitted to trial surveys on ITN use (see Appendix 5)<br>Half-life: 1.7 years for Olyset Plus and 2.4 years for Interceptor G2<br>Shape: 1.9 for Olyset Plus and 2.4 for Interceptor G2 | Weibull decay function with fixed half-life and shape parameters<br>Half-life: 3 years<br>Shape: 2 (assumed) |
| **Usage at distribution**<br>(Proportion of people sleeping under a net at night just after the net distribution) | 69% for Interceptor G2 arm and 75% for Olyset Plus (see Appendix 5) | 100% |
| **Insecticidal durability** | Linear decay between estimates for unwashed and washed ITNs in the EHT, assuming that washed nets represent nets after three years of use. For EHT trial arm *i* this corresponds to the following half-lives for deterrence, pre-prandial and post-prandial killing effects respectively:<br>$L_{\pi_i} = \frac{3}{1 - \pi_i^{unwashed} / \pi_i^{washed}}$ $L_{\phi_i} = \frac{3}{1 - \phi_i^{unwashed} / \phi_i^{washed}}$ | Estimates for unwashed nets are maintained throughout |
| | $L_{\xi_i} = \frac{3}{1 - \xi_i^{unwashed} / \xi_i^{washed}}$ | |
| **Activity rhythms** | Exposure in bed is 69% (see Table 2) | Exposure in bed is 100% |

### Reproducing *in silico* randomized control trial with a mathematical model of malaria transmission

Two randomized controlled trials conducted in Tanzania are considered in this analysis, Protopopoff et al. ^53,54^ and Mosha et al. ^12,13^ as they compare the efficacy of pyrethroid nets with new-generation nets including PBO and chlorfenapyr. They are reproduced *in silico* using the OpenMalaria simulation platform ^45^. This individual-based model accounts for the main specificities of malaria transmission dynamics in the human host (including immunity and superinfection) and the mosquito vector, and it has been described extensively elsewhere ^45^.

The RCTs are modelled by adapting OpenMalaria to the setting-specific conditions of each trial location, using parameters as indicated in Table 2. The mathematical model is adjusted to represent the control arms as well as the pre-intervention prevalence in the intervention arms, by calibrating three input parameters as explained below. RCT measurements on post-intervention prevalence are used for out-of-sample validation. The simulations are conducted with OpenMalaria v44 and the OpenMalariaUtilities R package, using a population of 10,000 individuals and 10 stochastic replicates for each scenario.

#### Entomological efficacy of ITNs in OpenMalaria

Within OpenMalaria, the effectiveness of ITNs is calculated using an *Anopheles* life-cycle model ^66,70^. In this model, the numbers of host-seeking, infected and infectious mosquitoes are simulated using deterministic difference-equations that quantify their survival probability across five stages of the feeding cycle (host-seeking, feeding, searching for a resting place, resting and ovipositing), when they attack protected or unprotected human hosts.

##### Pyrethroid-only ITNs

Pyrethroid-only ITNs are modelled as a “Generic Vector Intervention” with parameters adjusted to best match the control arm in each RCT, using the following procedure. For each trial, a grid of simulations with varying transmission intensity (EIR values between 10 and 200), pyrethroid-only nets entomological efficacy (pre-prandial killing effect between 0 and 1, deterrence and post-prandial killing effects fixed to 0 for identifiability reasons) and lag between rainfall and EIR to represent seasonality (1, 2, and 3-month lag) was created. The parameter set minimizing the least-square distance between simulated and observed prevalence for all measurements in the control arm was selected. The corresponding best-fitting seasonality lag and entomological efficacy were recorded. The EIR was then calibrated to best represent the pre-intervention prevalence, as detailed in Appendix 7.

In Protopopoff et al. ^54^, the observation at 28 months in the control arm was considered to be an outlier and it was decided not to be used for calibration - it is nonetheless displayed as a validation point.

For both trials, the best fit was obtained with a seasonality lag of 3 months. For Protopopoff et al. ^53^, the best fit was obtained with a pre-prandial killing effect of 0.05. For Mosha et al.^12^, the best fit was obtained with a pre-prandial killing effect of 0, although the value of 0.05 provided the second-best estimate. Therefore, for consistency, a value of 0.05 was used in both cases.

The final uncertainty range includes i) model stochasticity (10 seeds) and ii) uncertainty in initial transmission intensity resulting from profile likelihood methods ^69^ (see Appendix 7).

##### Next-generation ITNs

Next-generation ITNs are included in this model as the combination of two “Generic Vector Interventions” deployed simultaneously and to the same individuals:

- A “pyrethroid-only” intervention, represented with a pre-prandial killing effect of 0.05
- An intervention representing the additional impact of the net compared to the pyrethroid-only net, with the previously fitted values for *π, ϕ* and *ξ* taken as deterrence, pre-prandial and post-prandial killing effects respectively.

In Mosha et al. ^12,13^, where alpha-cypermethrin nets (Interceptor) were deployed in the control arm, the entomological efficacy of Interceptor G2 is informed from BIT103, BIT080 and Kibondo et al. ^51^ and the efficacy of Olyset Plus nets ITNs is informed from BIT103 only. In Protopopoff et al. ^53,54^, where permethrin nets (Olyset) were used in the control arm, BIT055 was used to inform the entomological efficacy of Olyset Plus relative to Olyset.

Uncertainty in the effect size is propagated by using three parameter sets from each EHT fitting, namely the posterior average and the upper and lower bounds of the 95% credible interval (because the relationship between all three parameters and the predicted effect size is monotonous, this corresponds to a conservative assumption).

The final uncertainty range includes i) model stochasticity (10 seeds), ii) uncertainty in initial transmission intensity resulting from profile likelihood methods ^69^ (see Appendix 7), iii) uncertainty from EHT fitting of entomological estimates (taking the envelope of the uncertainties due to EHT statistical fitting and EHT source data).

### Epidemiological effectiveness of ITNs in OpenMalaria

The OpenMalaria parameterisation includes various factors that influence ITN effectiveness beyond the sole entomological efficacy.

Firstly, the effectiveness of the ITNs depends on the proportion of individuals having access to and using them. In the model, the proportion of protected and unprotected hosts is informed by the proportion of individuals effectively using a net, inferred from RCT survey data on the proportion of individuals reporting using a net the night before the survey, as explained in Appendix 5.

Secondly, the durability of ITNs over the 3-year period between distribution campaigns is accounted for via two mechanisms, namely insecticidal durability and functional survival. Insecticidal durability is informed using the entomological estimates for 20x washed and unwashed nets. The estimates obtained for unwashed nets were used to represent the initial entomological efficacy. The estimates obtained for nets washed 20 times were used to represent the entomological efficacy after three years ^52^, assuming a linear decay over the 3-year time interval. Functional survival is included by reducing the effective ITN usage over time following a Weibull function with half-life and shape parameter fitted to RCT data on ITN usage as explained in Appendix 5.

Thirdly, because insecticide treated nets can only protect hosts that are bitten while in bed, published data on activity rhythms for both humans and mosquitoes from Tanzania (see Appendix 6 for details) are used to quantify the exposure to mosquito bites while in bed (denoted *ϵ*) ^21^. The corrected values for the deterrence, pre-prandial, and post-prandial killing effects thus become 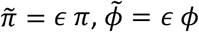 and 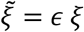.

#### Cascades of ITN effectiveness

The cascade of effectiveness relies on the concept of vectorial capacity, defined and calculated as explained previously.

The effectiveness cascade includes 5 different factors in this order: entomological efficacy, functional survival, imperfect usage at distribution, insecticidal durability and activity rhythms. For each of these factors, we compute a realistic estimate (derived previously to reproduce the RCT by Mosha et al. ^12,13^) and a perfect comparator corresponding to an ideal setting where there would be no loss of effectiveness due to the factor. For example, for ITN usage, realistic values from the trial are between 70 and 80%, while the perfect comparator would be 100%. All these estimates and their perfect comparator counterpart are detailed in Table 3.

To derive the effectiveness cascade, the vectorial capacity is first calculated considering the perfect comparator for each factor, and then re-calculated replacing sequentially each factor by its realistic value.

In order to propagate the fitting uncertainty of the entomological efficacy estimates, vectorial capacity is calculated for 1000 random samples from the posterior distribution for each EHT. The posterior mean and the 95% credible intervals in the vectorial capacity across the samples of all EHT for each net type were selected (posterior mean and 95% credible interval for each EHT are presented in Appendix 1).

The cascades presented in the main text are parameterized to represent the setting in Mosha et al. 2022; 2024), considering *Anopheles gambiae* vectors. Additionally, a dashboard is available to the reader to adapt the cascades to their own data.

## Data Availability

The transmission dynamics mathematical model is written in C++ and R and model code can be found here: https://github.com/SwissTPH/openmalaria, here: https://github.com/SwissTPH/r-openMalariaUtilities and here: https://github.com/SwissTPH/AnophelesModel. Data manipulation code, input parameters and processing code in R are available here: https://github.com/SwissTPH/ITNcascades.

## Acknowledgements

The authors want to thank Thomas Smith for helpful discussion over the course of the project and Rémi Turquier for visualization support.

Calculations were performed at the sciCORE (http://scicore.unibas.ch/) scientific computing core facility at the University of Basel.

This work was supported in whole or in part by the Bill & Melinda Gates Foundation [INV-030449 and INV-068864]. Under the grant conditions of the Foundation, a Creative Commons Attribution 4.0 Generic License has already been assigned to the Author Accepted Manuscript version that might arise from this submission.

The funders did not play any role in the study design, data collection and analysis, decision to publish, or preparation of the manuscript.

## Author contribution

This work was conceived by C.C, J.L., S.J.M. and E.P. Empirical data were produced by A.A., U.A.K., R.G.L., E.M., J.M., J.B.M., W.S.N., O.G.O., J.K.S. and S.J.M. C.C. performed the statistical analysis, with support from M.A. C.C. and J.L. performed the modelling simulations, with support from R.G. and M.G. C.C. performed the post-processing analysis, produced all figures and wrote the first draft. Expert guidance on analysis and interpretation were provided by S.J.M., N.C, A.R and E.P. All authors contributed comments and approved the final version of the manuscript.

## Ethics approval

Volunteers were recruited on written informed consent. Ethical approval from Ifakara Health Institute-Institutional Review Board and National Institute of Medical Research Tanzania were obtained respectively IHI/IRB/AMM/No:3–2021 and NIMRI/HQ/R.8c/Vol.I/908 for Kibondo, IHI/IRB/No: 29-2020 and NIMR/HQ/R.8a/Vol.IX/3521 for BIT055 and Odufuwa, IHI/IRB/No: 35-2021 and NIMR/HQ/R.8a/Vol.IX/3957 for BIT080, and IHI/IRB/No: 21-2023 and NIMR/HQ/R.8a/Vol.IX/4558 for BIT103.

## Competing interests

A.A., U.A.K., R.G.L., E.M., J.M., J.B.M., W.S.N., O.G.O., J.K.S. and S.J.M test vector control products for a range of manufacturers. The other authors declare no competing interests.

## Data availability

Data from experimental hut trials is available as follows. For Kibondo et al.^51^ and Odufuwa et al.^50^, the data is available in the original publications. For BIT103, BIT055 and BIT080, data is available as a supplementary material

## Notes

### Author Declarations

Volunteers were recruited on written informed consent. Institutional Review Board of Ifakara Health Institute and National Institute of Medical Research Tanzania gave ethical approval for this work.

